# Mild and moderate COVID-19 during Alpha, Delta and Omikron pandemic waves in urban Maputo, Mozambique, December 2020-March 2022: a population-based surveillance study

**DOI:** 10.1101/2023.12.22.23300474

**Authors:** Brecht Ingelbeen, Victória Cumbane, Ferão Mandlate, Barbara Barbé, Sheila Mercedes Nhachungue, Nilzio Cavele, Cremildo Manhica, Catildo Cubai, Neusa Maimuna Carlos Nguenha, Audrey Lacroix, Joachim Mariën, Anja de Weggheleire, Esther van Kleef, Philippe Selhorst, Marianne AB van der Sande, Martine Peeters, Marc-Alain Widdowson, Nalia Ismael, Ivalda Macicame

## Abstract

In sub-Saharan Africa, reported COVID-19 numbers have been lower than anticipated, even when considering populations’ younger age. The extent to which risk factors, established in industrialised countries, impact the risk of infection and of disease in populations in sub-Saharan Africa, remains unclear. We estimated the incidence of mild and moderate COVID-19 in urban Mozambique and analysed factors associated with infection and disease in a population-based surveillance study.

During December 2020-March 2022, households of a population cohort in Polana Caniço, Maputo, Mozambique, were contacted biweekly. Residents reporting any respiratory sign, anosmia, or ageusia, were asked to self-administer a nasal swab, for SARS-CoV-2 PCR testing. Of a subset of 1400 participants, dried blood spots were repeatedly collected three-monthly from finger pricks at home. Antibodies against SARS-CoV-2 spike glycoprotein and nucleocapsid protein were detected using an in-house developed multiplex antibody assay. We estimated the incidence of respiratory illness and COVID-19, and SARS-CoV-2 seroprevalence. We used Cox regression models, adjusting for age and sex, to identify factors associated with first symptomatic COVID-19 and with SARS-CoV-2 sero-conversion in the first six months.

During 11925 household visits in 1561 households, covering 6049 participants (median 21 years, 54.8% female, 7.3% disclosed HIV positive), 1895.9 person-years were followed up. Per 1000 person-years, 364.5 (95%CI 352.8-376.1) respiratory illness episodes of which 72.2 (95%CI 60.6-83.9) COVID-19 confirmed, were reported. Of 1412 participants, 2185 blood samples were tested (median 30.6 years, 55.2% female). Sero-prevalence rose from 4.8% (95%CI 1.1-8.6%) in December 2020 to 34.7% (95%CI 20.2-49.3%) in June 2021, when 3.0% were vaccinated. Increasing age (strong gradient in hazard ratio, HR, up to 15.70 in ≥70 year olds, 95%CI 3.74-65.97), leukaemia, chronic lung disease, hypertension, and overweight increased risk of COVID-19. We found no increased risk of COVID-19 in people with HIV or tuberculosis. Risk of COVID-19 was lower among residents in the lowest socio-economic quintile (HR 0.16, 95%CI 0.04-0.64), with no or limited handwashing facilities, and who shared bedrooms (HR 0.42, 95%CI 0.25-0.72). Older age also increased the risk of SARS-CoV-2 seroconversion (HR 1.57 in 60-69 year olds, 95%CI 1.03-2.39). We found no associations between SARS-CoV-2 infection risk and socio-economic, behavioural factors and comorbidities.

Active surveillance in an urban population cohort confirmed frequent COVID-19 underreporting, yet indicated that the large majority of cases were mild and non-febrile. In contrast to industrialised countries, deprivation did not increase the risk of infection nor disease.

## Introduction

Older age, deprivation, black ethnicity (compared to white) increased the risk of COVID-19 disease or death in different contexts (1–3). Younger age groups less frequently manifested symptoms or were hospitalised when infected, but also had lower infection rates (4,5). In sub-Saharan African countries, reported COVID-19 cases and deaths were lower than expected from the infection prevalence measured in sero-surveys and age-specific infection fatality (6,7). HIV increased the risk of COVID-19-related death (8), as did widely established risk factors as hypertension, diabetes, or chronic kidney of pulmonary disease (1). Higher COVID-19 incidence among deprived has been explained by inequalities in the ability to work remotely, and by higher secondary infection rates within (more crowded) households (9).

The extent to which non-pharmaceutical interventions had an impact on COVID-19 incidence in sub-Saharan African settings is still poorly understood. Other studies in Eastern Africa have shown a reduction in deaths from acute respiratory infections (10).

In Mozambique, 184219 COVID-19 cases and 2010 deaths were reported during 2020-21, yet excess deaths due to the pandemic have been estimated at 78100 (95%CI 54100-109000) (11). The excess mortality rate (139 per 100000, 95%CI 96-194) was comparable to the global all-age estimate (120 per 100000, 95%CI 113–129). COVID-19 vaccination started on 7 March 2021. By 8 September 2021, 5.0% of the Mozambican population received at least one dose of vaccine. By 8 March 2022, 40.2% did.

In Maputo City, the capital of Mozambique, in 2020-21, COVID-19 testing was centralised in two COVID-19 management facilities, where mild/moderate cases were unlikely to go. We provide a comprehensive description of the clinical range of mild and moderate COVID-19 in urban Maputo. We then estimate SARS-CoV-2 (infection) sero-prevalence, COVID-19 (disease) incidence rates, and analyse demographics, comorbidities, and exposures increasing the risk of infection and of disease.

## Methods

### Study design and population

Between December 2020 and March 2022, population based surveillance in urban Maputo consisted of two components: biweekly follow-up of households to record possible COVID-19 cases (acute respiratory symptom, fever, anosmia, or ageusia) and three-monthly sero-surveys to track SARS-CoV-2 antibodies in a subset of participants. Recruited households were embedded in the Health and Demographic Surveillance System of Polana Caniço, covering 15,393 residents. Participants had to be residents since ≥3 months, and included all ages.

At baseline, household-level demographics, socio-economic, water- and sanitation conditions, individual comorbidities, and behaviour potentially determining exposure were recorded in electronic questionnaires. Subsequently, households were visited or phoned every two weeks during one year to detect possible COVID-19 cases at the time of the visit or with symptom onset in the two weeks prior to the visit. Possible cases were asked to self-administer a nasal swab for SARS-CoV-2 PCR testing. Symptoms during the previous two weeks were recorded. If COVID-19 was confirmed, the case was followed up after 28 and 56 days to record clinical outcome.

For the repeated sero-survey, randomly selected participating household members from three age strata (0-17, 18-49, and ≥50 years) were visited every three months during one year to collect dried blood spots from a fingerprick and (from 31 March 2021 onwards) record prior COVID-19 vaccination.

### Laboratory procedures

Nasal swabs were self-administered at participants’ homes or assisted by interviewers in participants under 5 years of age, then transported in RNA Shield^™^ reagent. Real-time reverse-transcription PCR was performed within the same day at Instituto Nacional de Saúde, Marracuene, Mozambique.

Dried blood spots, containing 450μl of blood sampled from a fingerprick in six circles (each approximately 75ul) on dried blood spot filter paper (Whatman 903^™^ Protein Saver Card), were prepared for testing by punching two discs of 4mm diameter (corresponding to 40 μl of blood), and eluted overnight in 160 μL of hypertonic phosphate buffered saline-BSA (dilution 1:40, phosphate buffered saline-1 % BSA-0.15 % Tween, pH 7.4, Sigma-Aldrich). Before use in the immunoassay, eluted samples have been further diluted to 1:200 in hypertonic phosphate buffered saline-BSA, according to Mariën *et al* (12). We then used an in-house developed multiplex antibody assay for the detection of anti-SARS-CoV-2 IgG: we coupled recombinant large spike glycoprotein S1 and S2 subunit, receptor-binding domain (RBD), and nucleocapsid-protein (NP) antigens derived from SARS-CoV-2 at Sino Biological to maximum of 1.25□× □10^6 paramagnetic MAGPLEX COOH-microsphere beads from Luminex Corporation as antibody targets. 150□μl of beads and diluted sera were added to each well, incubated at room temperature, then washed with 200□μl/well of hypertonic phosphate buffered saline-BSA. Adding biotin-labelled anti-human secondary IgG and streptavidin-R-phycoerythrin conjugate, another 30 min incubation, samples were read on Luminex MagPix^™^ at Instituto Nacional de Saúde, Marracuene, Mozambique. For each antigen target, a cut-off value of antibody detection was estimated by adding 2.5 standard deviations to the average value of 42 negative control samples from residents of Maputo, collected prior to the pandemic and also spotted on filter paper. We used two citeria to determine seropositivity: (i) both RBD and NP above the cut-off, ensuring excellent specificity as demonstrated in (12), and (ii) RBD above the cut-off, similar to assays used in most other SARS-CoV-2 sero-surveys (5). After breakdown of the MagPix^™^ platform, serological testing for samples collected after August 2021 became unavailable.

### Data analysis

We estimated incidence rates of acute respiratory illness and of COVID-19 from respectively the number of possible cases and of confirmed COVID-19 cases, divided by the observation time. Because every household visit recorded possible cases with onset during two weeks before the visit, observation time consisted of the two weeks prior to each visit, or the time between visits if less than two weeks spanned between consecutive visits. We analysed clinical signs and symptoms associated to COVID-19, comparing confirmed cases to SARS-CoV-2 negative cases, adjusting for age using unconditional logistic regression.

To identify participant demographic, health, socio-economic and behavioural characteristics associated with COVID-19, we did a survival analysis fitting a Cox proportional hazards model with self-reported first confirmed COVID-19 as event variable, and observation time, censored after a first confirmed COVID-19 episode, as time variable, adjusting for age and sex.

We estimated infection- and vaccination-induced SARS-CoV-2 sero-prevalence, by age group, based on sero-survey samples collected up to 31 July 2021. Samples were SARS-CoV-2 sero-positive when antibodies against RBD and NP were detected, as proposed by a validation study of the assay (12). To ensure comparability to results with other sero-surveys, we analysed sero-prevalence based on antibodies against RBD only.

To identify participant characteristics associated with SARS-CoV-2 infection (including asymptomatic), similar to the above survival analysis of first symptomatic COVID-19, we fitted a Cox proportional hazards model to sero-survey participants with ≥ 2 samples collected up to 31 July 2021. Events were either a SARS-CoV-2 positive result following a negative result (sero-conversion) or an initial SARS-CoV-2 positive result. Time consisted of three months prior to each sero-survey, or the time between consecutive sero-surveys if less than three months spanned in-between, and was – in case of sero-conversion – censored at the midpoint between the last negative test and the subsequent positive test.

### Ethical considerations

The study protocol was approved by the Mozambican national health bio-ethics committee (517/CNBS/2020) and the Antwerp University Hospital ethics committee (B3002020000123). Study participants provided written informed consent at baseline for study participation, and again at the time of collecting a nasal swab or at the first sero-survey visit.

## Results

### Household surveillance of acute respiratory illness

Between 15 December 2020 and 31 March 2021, we conducted 11925 household visits in 1561 households, covering 6049 participants (Figure 1). Participants were median 21 years old (interquartile range, IQR, 11-38 years), 3315 (54.8%) were female, and 435 (7.3%) disclosed to be HIV positive. 2694 (55.6% of 4841 with recorded socio-economic status) did not complete primary education and 93 (1.9%) had higher education.

**Figure 1.**
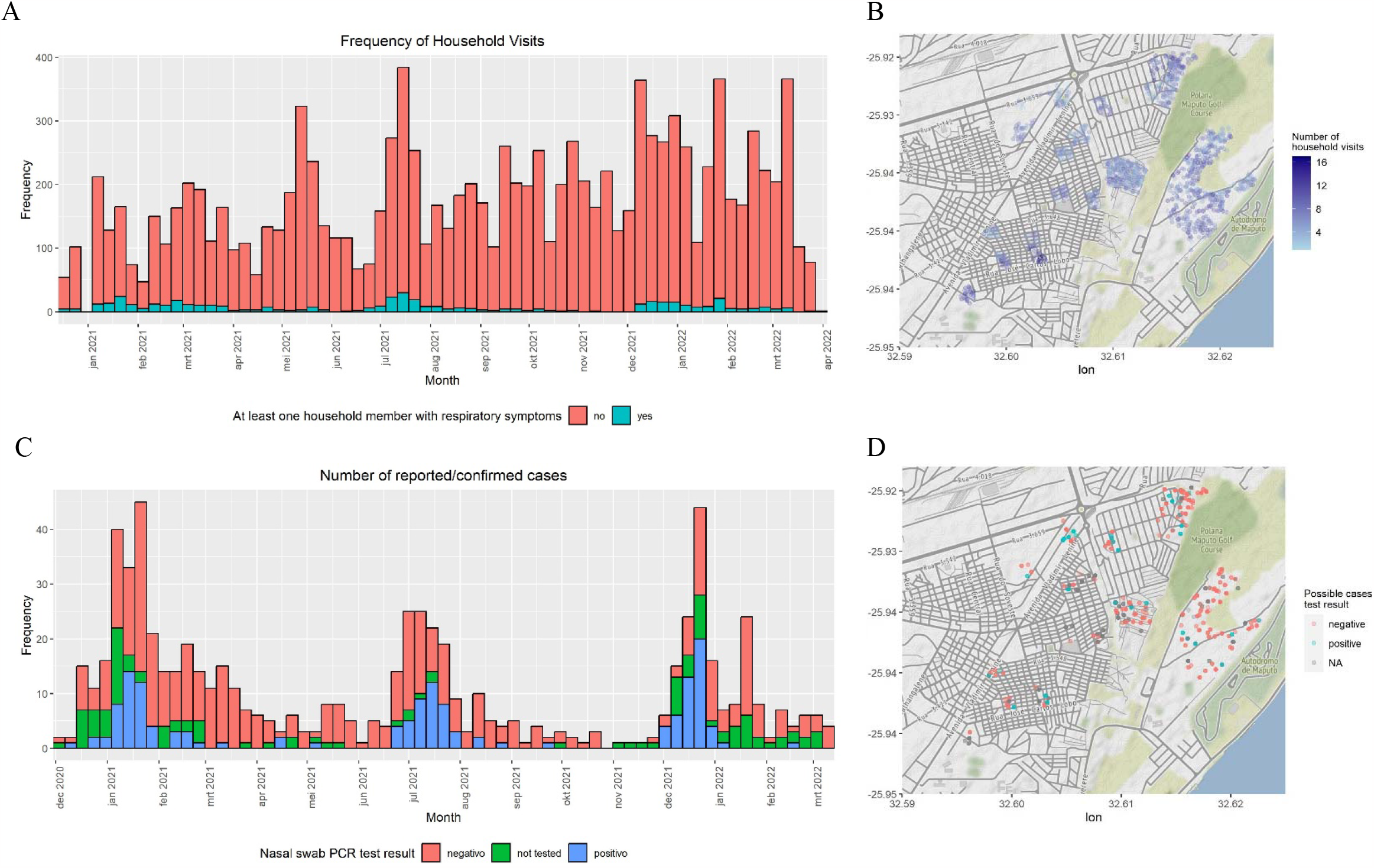
Household acute repiratory illness and COVID-19 surveillance. A. Weekly frequency of household visits, B. Geographical distribution of household visits in Maputo City, C. Weekly number of possible COVID-19 cases, stacked by SARS-CoV-2 PCR test result, D. Geographical distribution of possible COVID-19 cases by result.

### Respiratory illness and COVID-19 incidence rate

In the two weeks prior to the visits, 482 households reported at least one possible case in the household. The incidence rate of respiratory illness was 364.5 (95% CI 352.8-376.1) per 1000 person-years (py, 691 possible cases in 611 participants; 1895.9 py followed up). Of 579 possible cases, a nasal swab was collected and tested, median 5 days (IQR, 3-8 days) after symptom onset. SARS-CoV-2 was confirmed in 144 (24.9%) cases. The incidence rate of confirmed COVID-19 was 72.2 (95% CI 60.6 83.9) per 1000 py. Among participants under 18 years old, this was 25.9 (95%CI 15.0-36.8) per 1000 py; in 18-49 year olds it was 79.9 (95%CI 61.2-98.5) per 1000 py; in ≥50 year olds, it was 188.3 (95%CI 141.8-234.9) per 1000 py. SARS-CoV-2 positivity of tested possible cases peaked at 39% in January 2021, at 39% in July 2021, and at 55% in December 2021.

### Clinical signs and symptoms of mild and moderate COVID-19

Reported COVID-19 cases were median 36.4 years old (IQR 22.3-57.5 years) and 87 (60.4%) were female (Table 1). Compared to SARS-CoV-2 negative cases (median age 26.0 years, IQR 10.8-49.3, 55.8% female), COVID-19 cases had more frequently anosmia (age-adjusted odds ratio, aOR 2.36 95%CI 1.48-3.58), ageusia (aOR, 2.29 95%CI 1.45-3.58), loss of appetite (aOR 2.21 95%CI 1.37-3.56), and chills (aOR 1.78 95%CI 1.05-2.97). During the Omikron variant wave starting December 2021, the association with each of these symptoms disappeared (anosmia aOR 1.1 95%CI 0.46-2.56, ageusia 0.70 95%CI 2.29-1.62, loss of appetite 1.25 95%CI 0.52-2.96, chills 0.88 95%CI 0.28-2.49). Of 92 confirmed COVID-19 cases followed up after 28 days, of whom 54 again after 56 days, one (1.1%) died.

**Table 1.** Clinical signs and symptoms associated with SARS-CoV-2 confirmation among acute respiratory illness (possible COVID-19 cases) reported during December 2020-March 2022.

| Factors | Confirmed SARS-CoV-2 (N=148) |  | Negative SARS-CoV-2 (N=435) |  | Crude odds ratio |  | Age-adjusted odds ratio |  |
| --- | --- | --- | --- | --- | --- | --- | --- | --- |
|  | n | % | n | % | OR | 95%CI | aOR | 95% CI |
| Age*: 0-9 years | 7 | 4.9 | 90 | 21.5 | ref |  |  |  |
| 10-19 years | 18 | 12.5 | 79 | 18.9 | **2.93** | **1.21-7.87** |  |  |
| 20-29 years | 34 | 23.6 | 62 | 14.8 | **7.05** | **3.10-18.3** |  |  |
| 30-39 years | 18 | 12.5 | 48 | 11.5 | **4.82** | **1.95-13.2** |  |  |
| 40-49 years | 14 | 9.7 | 37 | 8.8 | **4.86** | **1.87-13.8** |  |  |
| 50-59 years | 24 | 16.7 | 46 | 11.0 | **6.71** | **2.82-17.9** |  |  |
| 60-69 years | 22 | 15.3 | 40 | 9.5 | **7.07** | **2.92-19.1** |  |  |
| 70+ years | 7 | 4.9 | 17 | 4.1 | **5.29** | **1.62-17.4** |  |  |
| Sex*: Female | 87 | 60.4 | 234 | 55.8 | 1.21 | 0.82-1.78 | 1.19 | 0.81-1.77 |
| Clinical signs/symptoms |  |  |  |  |  |  |  |  |
| Cough | 117 | 89.3 | 326 | 86.0 | 1.36 | 0.75-2.63 | 1.52 | 0.82-2.95 |
| Headache | 81 | 61.8 | 217 | 57.3 | 1.21 | 0.81-1.82 | 1.14 | 0.75-1.73 |
| Rhinorrhoea | 69 | 52.7 | 221 | 58.3 | 0.80 | 0.53-1.19 | 0.86 | 0.57-1.29 |
| Throat pain | 54 | 41.2 | 131 | 34.6 | 1.33 | 0.88-1.99 | 1.22 | 0.80-1.84 |
| Ageusia | 47 | 35.9 | 69 | 18.2 | **2.51** | **1.61-3.91** | **2.29** | **1.45-3.58** |
| Oxygen saturation <95% | 16/48 | 33.3 | 36/151 | 23.8 | 1.60 | 0.78-3.22 | 1.54 | 0.74-3.14 |
| Anosmia | 43 | 32.8 | 63 | 16.6 | **2.45** | **1.55-3.85** | **2.36** | **1.48-3.75** |
| Fatigue | 41 | 31.3 | 95 | 25.1 | 1.36 | 0.88-2.10 | 1.26 | 0.80-1.96 |
| Loss of appetite | 39 | 29.8 | 62 | 16.4 | **2.19** | **1.37-3.48** | **2.21** | **1.37-3.56** |
| Fever | 33 | 25.2 | 96 | 25.3 | 0.99 | 0.62-1.56 | 1.10 | 0.68-1.75 |
| Chills | 29 | 22.1 | 50 | 13.2 | **1.87** | **1.12-3.10** | **1.78** | **1.05-2.97** |
| Joint pain | 29 | 22.1 | 49 | 12.9 | **1.91** | **1.14-3.17** | 1.59 | 0.90-2.59 |
| Chest pain | 25 | 19.1 | 60 | 15.8 | 1.25 | 0.74-2.08 | 1.10 | 0.64-1.85 |
| Myalgia | 24 | 18.3 | 49 | 12.9 | 1.51 | 0.87-2.56 | 1.24 | 0.70-2.13 |
| Nausea | 10 | 7.6 | 23 | 6.1 | 1.28 | 0.57-2.69 | 1.29 | 0.57-2.77 |
| Dyspnoe | 9 | 6.9 | 36 | 9.5 | 0.70 | 0.31-1.44 | 0.62 | 0.27-1.31 |
| Diarrhoea | 8 | 6.1 | 26 | 6.9 | 0.88 | 0.37-1.92 | 0.86 | 0.35-1.89 |
| Vomit | 7 | 5.3 | 19 | 5.0 | 1.07 | 0.41-2.50 | 1.32 | 0.49-3.23 |
| Rash | 3 | 2.3 | 3 | 0.8 | 2.94 | 0.54-16.0 | 2.98 | 0.54-16.4 |
| Nose bleeding | 1 | 0.8 | 3 | 0.8 | 0.95 | 0.05-7.5 | 0.99 | 0.05-7.94 |
| Change of consciousness | 0 | 0.0 | 3 | 0.8 |  |  |  |  |

\* age and sex missing of 16 possible cases, clinical signs and symptoms missing of 69 possible cases.

### Characteristics associated with (symptomatic) COVID-19

Increasing age (in ≥70 year olds hazard ratio (HR) 15.70, 95%CI 3.74-65.97) and several reported comorbidities increased the risk of symptomatic COVID-19: leukaemia, chronic lung disease, overweight, underweight, diabetes, chronic heart disease, and hypertension (Table 2). We found no increased risk of COVID-19 in people with HIV (HR 0.80, 95%CI 0.37-1.76) or with (a history of) tuberculosis (HR 0.93, 95%CI 0.33-2.58). The risk of COVID-19 was lower in people belonging to the lowest socio-economic quintile (HR 0.16, 95%CI 0.04-0.64), in households with no or limited handwashing facilities, and in households where bedrooms were shared with 3 or more household members (HR 0.42, 95%CI 0.25-0.72).

**Table 2.**
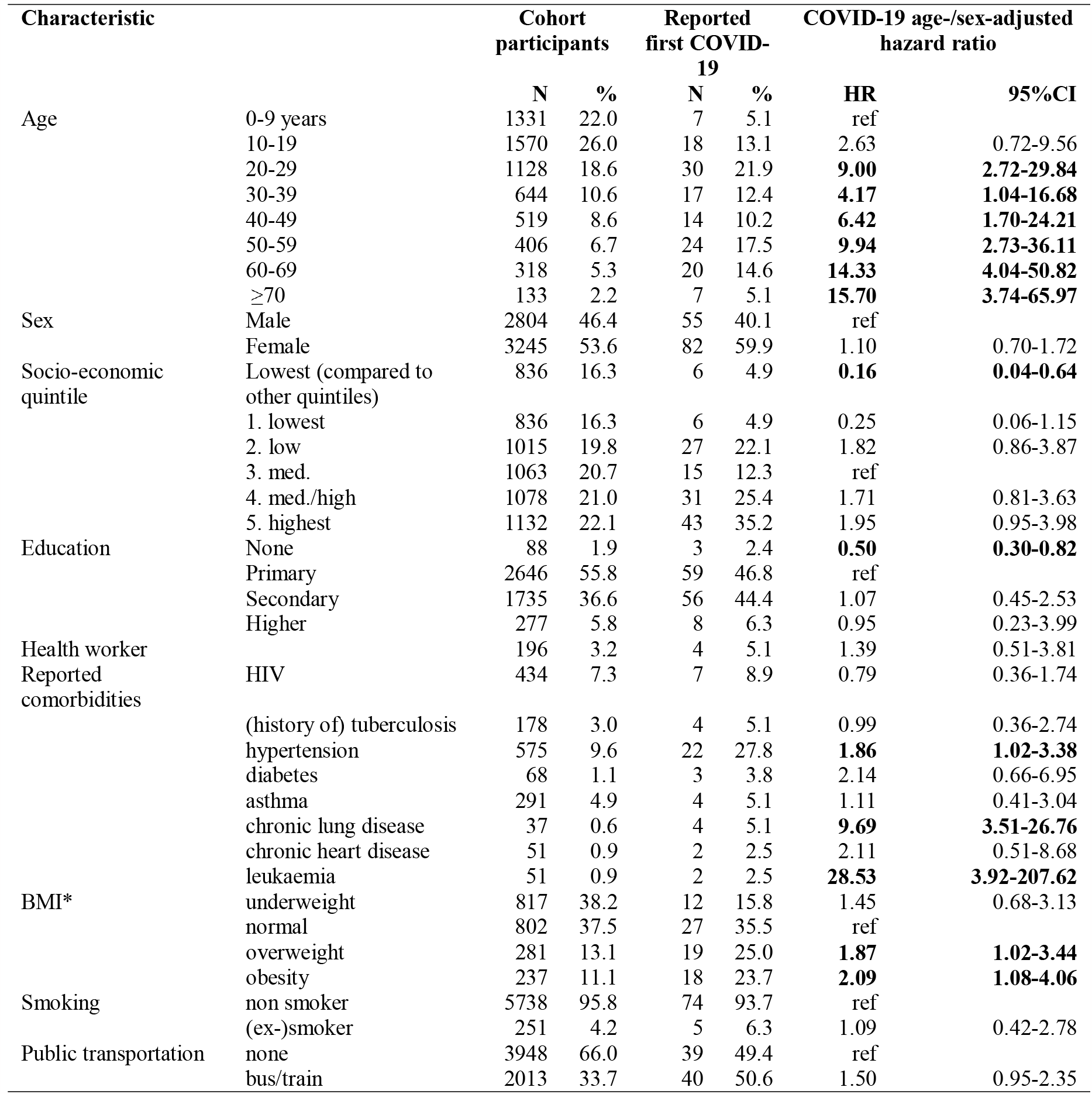
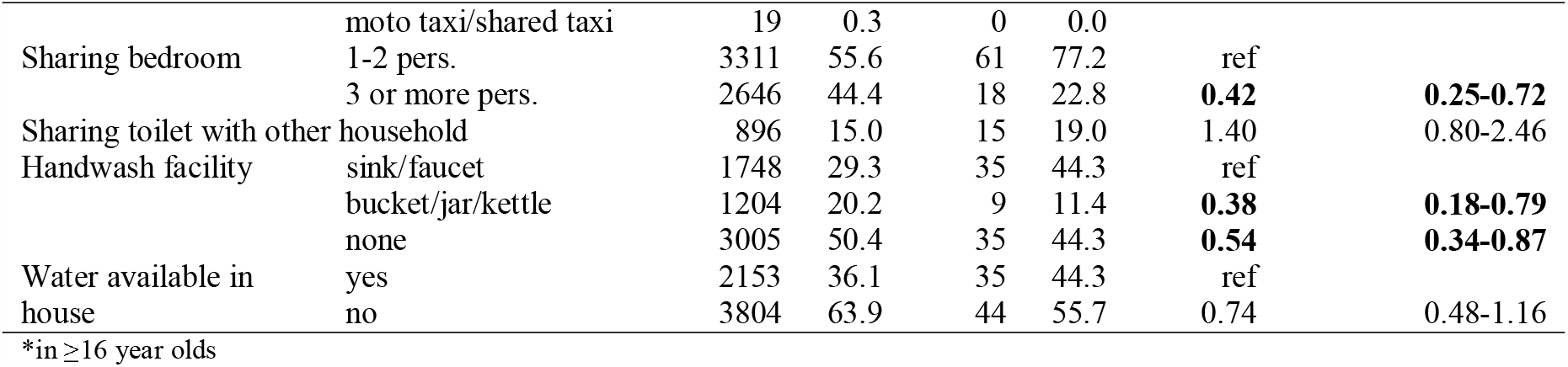
Demographic, socio-economic and behavioural characteristics associated with first confirmed COVID-19 among among acute respiratory illness (possible COVID-19 cases) reported in population-based surveillance during December 2020-March 2022, using a Cox regression model adjusting for age and sex.

### Infection-induced SARS-CoV-2 sero-prevalence

2185 samples collected until 31 July 2021 of 1412 sero-survey participants (median age 30.6 years, IQR 13.7-57.6; 38.2% ≥50 years old; 55.2% female) were tested. 301 participants (21.3%) tested positive (antibodies against RBD and NP) at least once. 34 (45.3%) out of 75 with a test subsequent to positive test, seroreverted.

Crude sero-prevalence increased from 4.8% (95%CI 1.1-8.6) in December 2020 to 34.2% (95%CI 23.4-45.1) in June 2021, when 2.7% of participants were vaccinated with at least one dose of vaccine (Figure 2). Sero-prevalence increased strongest in ≥50 year olds, peaking at 51.6% (95%CI 34.0-69.2) in June 2021, when 3.2% was vaccinated, yet declined to 41.6% (95%CI 26.5-56.7) in July 2021 when 10.9% was vaccinated.

**Figure 2.**
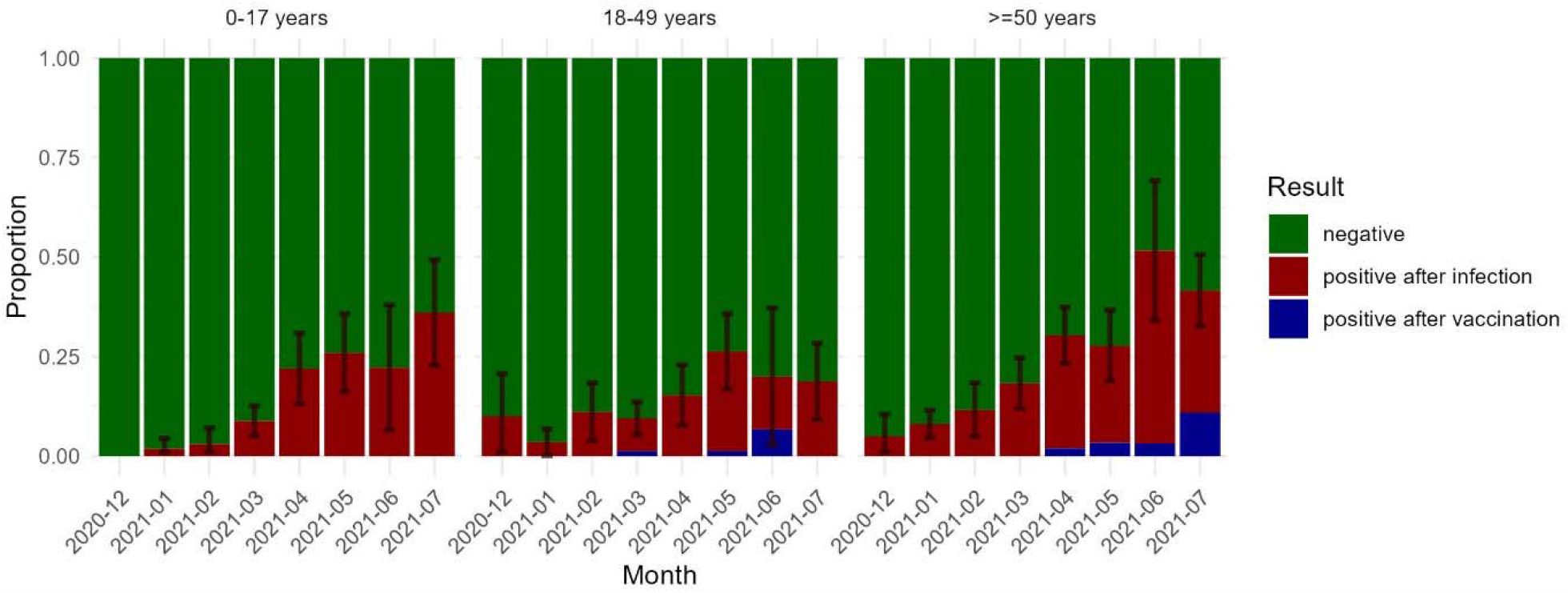
Infection- and vaccine-induced SARS-CoV-2 sero-prevalence by age group, December 2020 - July 2021. N_0-17 years_=647, N_18-49 years_=612, N_≥50 years_=882.

Crude sero-prevalence based on antibodies against RBD only was higher, rising from 10.5% (95%CI 5.1-15.9%) in December 2020 to 46.5% (95%CI 36.7-56.3) in July 2021, without decrease in sero-prevalence from June to July 2021 (Supplementary table in Appendix).

### Characteristics associated with SARS-CoV-2 infection

Older age increased the risk of SARS-CoV-2 infection (HR 60-69 versus 0-9 years 1.57, 95%CI 1.03-2.39, Table 3). We found no association between SARS-CoV-2 infection risk and socio-economic, behavioural factors and comorbidities.

**Table 3.**
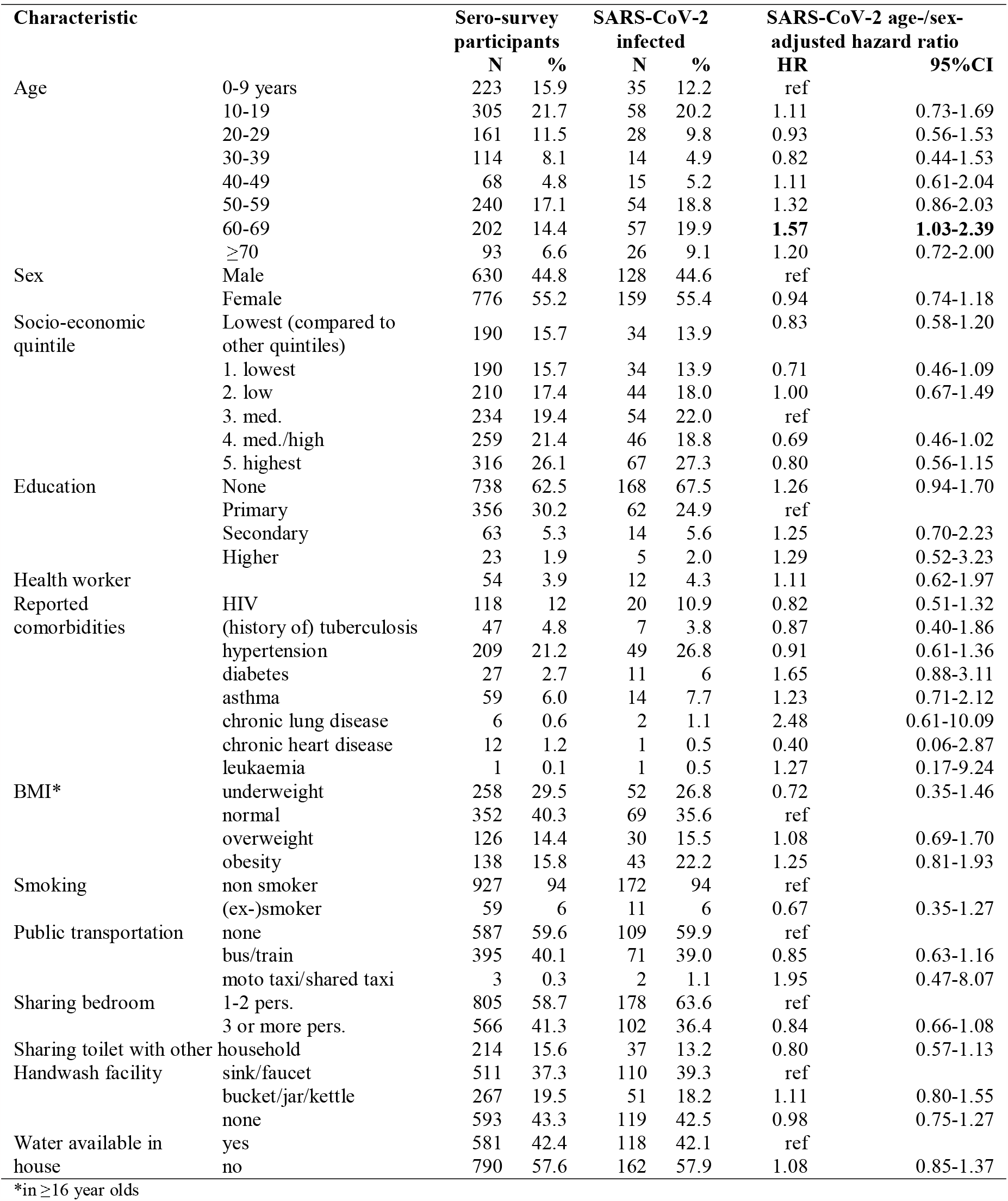
Demographic, socio-economic and behavioural characteristics associated with SARS-CoV-2 infection (including asymptomatic) among sero-survey participants with ≥ 2 samples tested during December 2020-July 2021, using a Cox regression model adjusting for age and sex.

## Discussion

Population-based COVID-19 surveillance in an urban population cohort in Mozambique confirmed that even during the acute phase of the pandemic the large majority of SARS-CoV-2 infections were asymptomatic, (symptomatic) COVID-19 cases were mild, and three in four were non-febrile.

Three COVID-19 peaks were distinct and died out after a few weeks, while other respiratory illness (SARS-CoV-2 negative) continued to be reported throughout 2021. Sudden drops in COVID-19 incidence after peaks while respiratory pathogens other than SARS-CoV-2 continue to circulate, indicate that newly introduced COVID-19 variants (Alpha, Delta, Omikron) could quickly spread despite non-pharmaceutical interventions. Nonetheless, transmission slowed when infection-induced seroprevalence (e.g., 11.2% in March 2021) was still lower than the herd immunity threshold anticipated from the Alpha variants’ reported transmissibility (13).

Surveillance started in December 2020 when the Beta variant had spread for 3 months through neighbouring South Africa, and two weeks before a surge in cases in Mozambique. Infection-induced seroprevalence of 4.8% in December 2020 was lower than the corrected pooled 16.2% reported in other African countries that month (5). Limited spread of wild type virus and the Beta variant during the first year of the pandemic – contrasting to neighbouring South Africa and Eswatini – could result from limited seeding through imported cases, limited mobility within the city and country, and longer maintained non-pharmaceutical interventions compared to its neighbours (14). The eventual surge in cases in January 2021 followed relaxed non-pharmaceutical interventions for the end-of-year holidays coinciding with the presumed introduction of the Alpha variant with increased transmissibility (13).

SARS-CoV-2 sero-prevalence rose to 34.7% in June 2021, still far below the pooled 76% infection-induced seroprevalence reported in other African countries. Only a fraction of that difference in sero-prevalence can be explained by serological test specificity. Our analysis of SARS-CoV-2 sero-positivity comparing several SARS-CoV-2 antigen targets demonstrated a difference of up to 12%, thus cannot explain the twice higher sero-prevalence in other sero-surveys in sub-Saharan Africa. A diagnostic performance study of the serological immunofluorescence assay supported the use of RBD and NP for IgG detection (12). A study using the same assay however showed decreasing sero-prevalence as a result of waning NP-specific IgG after three months (15). This could explain the decreasing sero-prevalence observed in July 2021.

The observed effect of age, obesity, and chronic conditions on the risk of mild disease was similar to the effect on risk of severe disease or death reported elsewhere (1,16). While HIV and (history of) tuberculosis have - been associated with COVID-19-related death (8), we observed no increased risk of infection nor of disease. Also, deprivation, increasing the risk of infection, disease and severity in several settings (1,2,17), did not affect the risk of SARS-CoV-2 infection in this population of Maputo city. Several indicators of deprivation, such as belonging to the lowest wealth quintile, no education, absence of handwash facilities, were even associated with a lower risk of COVID-19.

Among cases of respiratory illness, only anosmia, ageusia, loss of appetite and chills increased the probability of COVID-19, yet none of those symptoms was reported by more than a third of cases and the association disappeared in cases from December 2021 onwards, presumably Omikron cases. Symptoms’ poor predictive value, in combination with continued reporting of other respiratory illness, hampers diagnosis of mild/moderate COVID-19 on clinical grounds alone (18).

## Data Availability

Pseudonymized data supporting the findings of this study/publication are retained at the Institute of Tropical Medicine, Antwerp and can be made available after approval of a motivated and written request to. Study protocol, data dictionaries, scripts for conducting the analysis, and anonymized data, without geo-located or other data that could allow identification, are available on https://github.com/ingelbeen/africover-git

## Transparency declaration

All authors declare no competing interests. The work was funded by a European & Developing Countries Clinical Trials Partnership (EDCTP) project (RIA2020EF-3031). The funders had no role in study design, data collection and analysis, decision to publish, or preparation of the manuscript.

## Acknowledgements

We thank participants of the Polana Caniço Health and Demographic Surveillance System, interviewers, teams involved in laboratory testing of samples (Claudia Machume, Gercio Cuamba, Caro Van Geel), data management (Alberto Machaze, Eben Matavele, Harry van Loen), and study monitors (Dimpall Asmucrai, Carolien Hoof).

## Appendix

**Supplementary Table 1.** Infection- and vaccine-induced SARS-CoV-2 seroprevalence based on detection of antibodies against different SARS-CoV-2 antigens

| month | antibody detection | RBP and NP positive |  |  | two out of RBD, NP and S1S2 positive |  |  | RBD positive (regardless of S1S2 and NP) |  |  |
| --- | --- | --- | --- | --- | --- | --- | --- | --- | --- | --- |
|  |  | n | prop | 95%CI | n | prop | 95%CI | n | prop | 95%CI |
| 2020-12 | negative | 118 | 0.952 | 0.914 0.989 | 112 | 0.903 | 0.851 0.955 | 111 | 0.895 | 0.841 0.949 |
| 2020-12 | positive after infection | 6 | 0.048 | 0.011 0.086 | 12 | 0.097 | 0.045 0.149 | 13 | 0.105 | 0.051 0.159 |
| 2021-01 | negative | 433 | 0.945 | 0.925 0.966 | 416 | 0.908 | 0.882 0.935 | 399 | 0.871 | 0.840 0.902 |
| 2021-01 | positive after infection | 25 | 0.055 | 0.034 0.075 | 42 | 0.092 | 0.065 0.118 | 59 | 0.129 | 0.098 0.160 |
| 2021-02 | negative | 205 | 0.911 | 0.874 0.948 | 195 | 0.867 | 0.822 0.911 | 192 | 0.853 | 0.807 0.900 |
| 2021-02 | positive after infection | 20 | 0.089 | 0.052 0.126 | 30 | 0.133 | 0.089 0.178 | 33 | 0.147 | 0.100 0.193 |
| 2021-03 | negative | 465 | 0.884 | 0.857 0.911 | 447 | 0.850 | 0.819 0.880 | 436 | 0.829 | 0.797 0.861 |
| 2021-03 | positive after infection | 59 | 0.112 | 0.085 0.139 | 77 | 0.146 | 0.116 0.177 | 88 | 0.167 | 0.135 0.199 |
| 2021-03 | positive after vaccination | 2 | 0.004 | -0.001 0.009 | 2 | 0.004 | -0.001 0.009 | 2 | 0.004 | -0.001 0.009 |
| 2021-04 | negative | 244 | 0.758 | 0.711 0.805 | 238 | 0.739 | 0.691 0.787 | 212 | 0.658 | 0.607 0.710 |
| 2021-04 | positive after infection | 75 | 0.233 | 0.187 0.279 | 81 | 0.252 | 0.204 0.299 | 107 | 0.332 | 0.281 0.384 |
| 2021-04 | positive after vaccination | 3 | 0.009 | -0.001 0.020 | 3 | 0.009 | -0.001 0.020 | 3 | 0.009 | -0.001 0.020 |
| 2021-05 | negative | 181 | 0.733 | 0.678 0.788 | 179 | 0.725 | 0.669 0.780 | 163 | 0.660 | 0.601 0.719 |
| 2021-05 | positive after infection | 62 | 0.251 | 0.197 0.305 | 64 | 0.259 | 0.204 0.314 | 79 | 0.320 | 0.262 0.378 |
| 2021-05 | positive after vaccination | 4 | 0.016 | 0.000 0.032 | 4 | 0.016 | 0.000 0.032 | 5 | 0.020 | 0.003 0.038 |
| 2021-06 | negative | 48 | 0.658 | 0.549 0.766 | 46 | 0.630 | 0.519 0.741 | 42 | 0.575 | 0.462 0.689 |
| 2021-06 | positive after infection | 23 | 0.315 | 0.209 0.422 | 25 | 0.342 | 0.234 0.451 | 29 | 0.397 | 0.285 0.510 |
| 2021-06 | positive after vaccination | 2 | 0.027 | -0.010 0.065 | 2 | 0.027 | -0.010 0.065 | 2 | 0.027 | -0.010 0.065 |
| 2021-07 | negative | 143 | 0.665 | 0.602 0.728 | 143 | 0.665 | 0.602 0.728 | 115 | 0.535 | 0.468 0.602 |
| 2021-07 | positive after infection | 61 | 0.284 | 0.223 0.344 | 61 | 0.284 | 0.223 0.344 | 86 | 0.400 | 0.335 0.465 |
| 2021-07 | positive after vaccination | 11 | 0.051 | 0.022 0.081 | 11 | 0.051 | 0.022 0.081 | 14 | 0.065 | 0.032 0.098 |

RBD= receptor-binding domain (of the spike protein), NP=nucleocapsid protein, S1S2=spike glycoprotein S1 and S2 subunit

